# Comparison of Monkeypox disease knowledge and perception among the healthcare workers versus the general population during the first month of the WHO emerging infectious disease alert

**DOI:** 10.1101/2022.10.17.22281183

**Authors:** Mohamad-Hani Temsah, Fadi Aljamaan, Shuliweeh Alenezi, Noura Abouammoh, Khalid Alhasan, Shereen A. Dasuqi, Ali Alhaboob, Mohammed A. Hamad, Rabih Halwani, Abdulkarim Alrabiaah, Sarah Al-Subaie, Fatimah Al-Shahrani, Fahad AlZamil, Ziad A Memish, Mazin Barry, Jaffar A. Al-Tawfiq

## Abstract

**Background:** Monkeypox disease (MPD) recently re-emerged in May 2022 and caused international outbreaks in multiple non-endemic countries. This study aimed to assess the Saudi Arabian public and healthcare workers (HCWs) knowledge about MPD and their information-seeking attitudes before any cases were yet reported in Saudi Arabia.

**Methods:** This online survey of HCWs and the public in the Kingdom of Saudi Arabia (KSA) was conducted from May 27 to June 5, 2022. The survey tool was adopted from our published research on COVID-19 with modifications related to the new MPD outbreak, which was validated for content, language, and consistency. Participants were invited by convenience sampling techniques through various social media platforms (i.e., Twitter and WhatsApp groups) and email lists.

Variables surveyed included participants’ sociodemographic and job-related characteristics, COVID-19 infection status, and advocacy for MPD vaccination. Multiple questions assessing the participants’ knowledge related to MPD and (MPV) in terms of transmission, vaccination information by HCWs, and required isolation precautions. Finally, a Generalized Anxiety Disorder (GAD7) score was calculated. We then assessed the independent variables associated with participants’ attitudes to seek more information about MPD, and those associated with knowledge scores.

**Results:** A total of 1546 participants completed the public survey, and 61.3% showed interest in seeking more information about MPD. Of the participants, 48.7% knew that MPD could be transmitted before skin blisters appear, and 62.7% inferred that skin lesions are infectious. Only 38.1% inferred that touching contaminated surfaces, and 46.5% knew that sexual contact is a mode of transmission. 56.6% falsely believed the old smallpox vaccine is not effective against MPD.

Public participants’ overall mean knowledge score of MPD was 4.88 of 9 points. In contrast, the mean score of the knowledge of the 1130 HCWs was 14.4 of 28. Among HCWs, 28.3% correctly answered that the Jynneos vaccine has activity against MPD and 79.7% incorrectly answered that VARIVAX, a chickenpox vaccine, is effective against MPD. In addition, 74.2% of HCWs perceived the need to seek more information about MPD. Male HCWs had significantly lower mean knowledge scores compared to females. Physicians and HCWs’ self-rated high awareness of MPD correlated positively and significantly with their knowledge score.

**Conclusion:** In this study, the general public and HCWs had a moderate level of knowledge about MPD. The knowledge gaps among HCWs were evident in the clinical presentation of patients and vaccinations. Both groups reported a desire to seek more information about MPD, but this did not correlate with knowledge scores. It is important to have further education and intensification of campaigns to enhance awareness of MPD. It is also crucial to have further studies evaluate the knowledge of both groups over time.

## Introduction

Monkeypox disease MPD recently re-emerged in May 2022 and caused international outbreaks in multiple non-endemic countries. As of July 13, 2022, a total of 10,845 laboratory-confirmed cases of MPD were reported in 59 countries with no historical report of the disease before[1]. Monkeypox virus (MPV) is a DNA virus that is a member of the genus Orthopoxvirus and is closely related to smallpox which caused more than 300 million fatalities during the twentieth century, but was ultimately eradicated in 1980 by global vaccination efforts led by the World Health Organization (WHO), which also had efficacy against MPV[2]. However, with the cessation of smallpox vaccination, relative partial immunity against MPV has waned and may have contributed to its re-emergence in endemic African countries[3].

MPD, originally a zoonotic disease, is transmitted through direct contact with infected animals’ blood, bodily fluids, cutaneous/mucosal lesions, or indirect transmission through eating inadequately cooked meat or other animal products. Human-to-human transmission has been described for MPD but in a very limited nature and usually in endemic areas of Africa[4]. Close contact with human respiratory secretions (usually prolonged face-to-face contact), skin lesions, or recently contaminated objects, put healthcare workers (HCWs), household members, and other close contacts of infected individuals at risk [5]. Although the prospect of sexual transmission was not wholly established, four monkeypox-positive cases in Italy raised that possibility as the viral DNA was found in their seminal fluid samples[6]. Another study supported the virus shedding in body fluids such as saliva, semen, urine, and feces[7].

With the recent emergence of cases in non-endemic countries, the initial WHO assessment did not consider MPD a Public Health Emergency of International Concern (PHEIC) [8]. However, in a subsequent assessment, the WHO declared MPD as a PHEIC[9,10]. In newly affected countries, this is the first time that most cases have been confirmed among men who have had recent sexual contact with a new or multiple male partners, and clinical presentation has been reported with atypical presentations, including anogenital and mucosal lesions[4]. MPD has been historically described mainly in the Sub-Saharan African countries, recently reported in multiple countries in outbreak fashion triggers researchers to. Assessment of the knowledge is critical to perceive the public and HCWs’ response to alerts, especially of infectious nature, in order to deliver high-quality care by (HCWs) and ensure both HCWs’ and the public’s compliance with healthy practices to prevent disease acquisition and spread.

This study aimed to assess the Saudi Arabian public and HCWs’ knowledge about MPD and their information-seeking attitude during the first month of the WHO alert about MPD, before any cases were yet reported in the Kingdom of Saudi Arabia (KSA), as the first MPD was reported by the Ministry of Health (MOH) on July 14, 2022[11].

## Methods

### Data Collection

An online survey of HCWs and the public in KSA was conducted from May 27 to June 5, 2022. Participants were invited by convenience sampling techniques through various social media platforms (Twitter and WhatsApp groups) and email lists. Participants were invited to complete the online survey through the SurveyMonkey© platform, with each response allowed once from each unique IP address to ensure single entries. The first page of the survey included the IRB approval, consent of participation explained the study research objectives, and assured confidentiality.

The survey tool was adopted from our published research on COVID-19 with modifications related to the new MPD outbreak [12–18]. The final version was tested for content validity by our assigned research experts and piloted among ten HCWs for clarity and consistency. Modifications were implemented based on the experts’ recommendations. The research team approved the final version of the survey for language accuracy, clarity, and content validity.

Variables surveyed included participants’ sociodemographic and job-related characteristics, COVID-19 infection status, and advocacy for MPD vaccination. Multiple questions assessing the participants’ knowledge related to MPD and (MPV) in terms of transmission, vaccination information by HCWs, and required isolation precautions. Finally, Generalized Anxiety Disorder (GAD7) [19,20], a self-reported, 7-item validated scale, was used to measure anxiety. We then assessed the independent variables associated with participants’ attitudes to seek more information about MPD, and the variables associated with knowledge score.

#### 1.1 Ethical Approval

Ethical approval was granted by the institutional review board (IRB) at King Saud University (22/0416/IRB).

#### 1.2 Statistical analysis

Means and standard deviations were used to describe continuous variables, frequencies, and percentages for categorically measured variables. The histogram and the Kolmogorov-Smirnov test were applied to test the assumption of normality, and Levene’s test was used to test the homogeneity of variance statistical assumption. Cronbach’s alpha test was used to assess the internal consistency of the measured questionnaires. The Multivariate Binary Logistic Regression Analysis was used to assess the variables’ independent correlation. The association between predictors with the categorically measured variables was assessed with multivariate Logistic Binary regression analysis which was expressed with adjusted Odds Ratio (OR) with their associated 95% confidence intervals. While the beta coefficient was used to assess the variables’ independent association with continuous variables. The SPSS IBM statistical analysis program was used for statistical data analysis. The statistical Alpha significance level was considered at 0.050 level.

## Results

### Publics’ Monkeypox knowledge assessment questions

A total of 1546 participants completed the public survey. Table 1 (Appendix) displays the details of their MPD knowledge assessment questions which were assessed with seven questions. 48.7% knew that it could be transmitted before skin blisters appear, and 46.5% could be transmitted through sexual contact. Regarding MPD treatment, 47.3% knew that there are effective antiviral medications, while 48% falsely answered there are not 56.6% falsely believed the old smallpox vaccine is not effective against MPD, while 36.4% knew it is. Due to the convergence of names and possible confusion, we assessed the participants’ perception if the chickenpox vaccine might be effective against MPD; 14.7% did not think it is, 64.7% were unsure, and 20.6% falsely thought it is.

**Table 1:** Multivariate Binary Logistic Regression Analysis of respondents’ odds of public High MPD Knowledge score.

|  | Multivariate adjusted Odds Ratio (OR) | 95% C.I. for (OR) |  | p-value |
| --- | --- | --- | --- | --- |
|  |  | Lower | Upper |  |
| **Age group** | 0.975 | 0.874 | 1.087 | 0.646 |
| **Educational Level : University degree or higher** | 1.358 | 1.118 | 1.649 | 0.002 |
| **Employed** | 1.172 | 0.935 | 1.467 | 0.168 |
| **Previously developed COVID-19** | 0.816 | 0.660 | 1.008 | 0.060 |
| **Self and family worry about contracting Monkeypox infection** | 1.843 | 1.486 | 2.286 | <0.001 |
| **Source of Monkeypox Information: MOH** | 2.274 | 1.772 | 2.919 | <0.001 |
| **Source of Monkeypox Information: CDC & WHO websites** | 1.583 | 1.215 | 2.064 | 0.001 |
| **Source of Monkeypox Information: social media** | 1.735 | 1.332 | 2.262 | <0.001 |
| **Source of Monkeypox Information: Other internet sources** | 1.757 | 1.001 | 3.082 | 0.049 |
| **Monthly Household Income** | 1.064 | 0.990 | 1.143 | 0.094 |

**DV= High MPD Knowledge score**

Regarding MPD transmission modes: 62.7% of the public correctly inferred that pimples are infective, 57.6 % correctly inferred that respiratory secretions are also, while only 46.5% correctly inferred that sexual contact and intimate relationships can be modes for transmission. However, only 38.1% had correctly inferred that touching contaminated surfaces with (MPV) is infective, and 14.4% incorrectly inferred that contaminated foods & drinks are possible modes of transmission. 3.4% believed other modes of transmission might exist (Figure 1).

**Figure 1:**
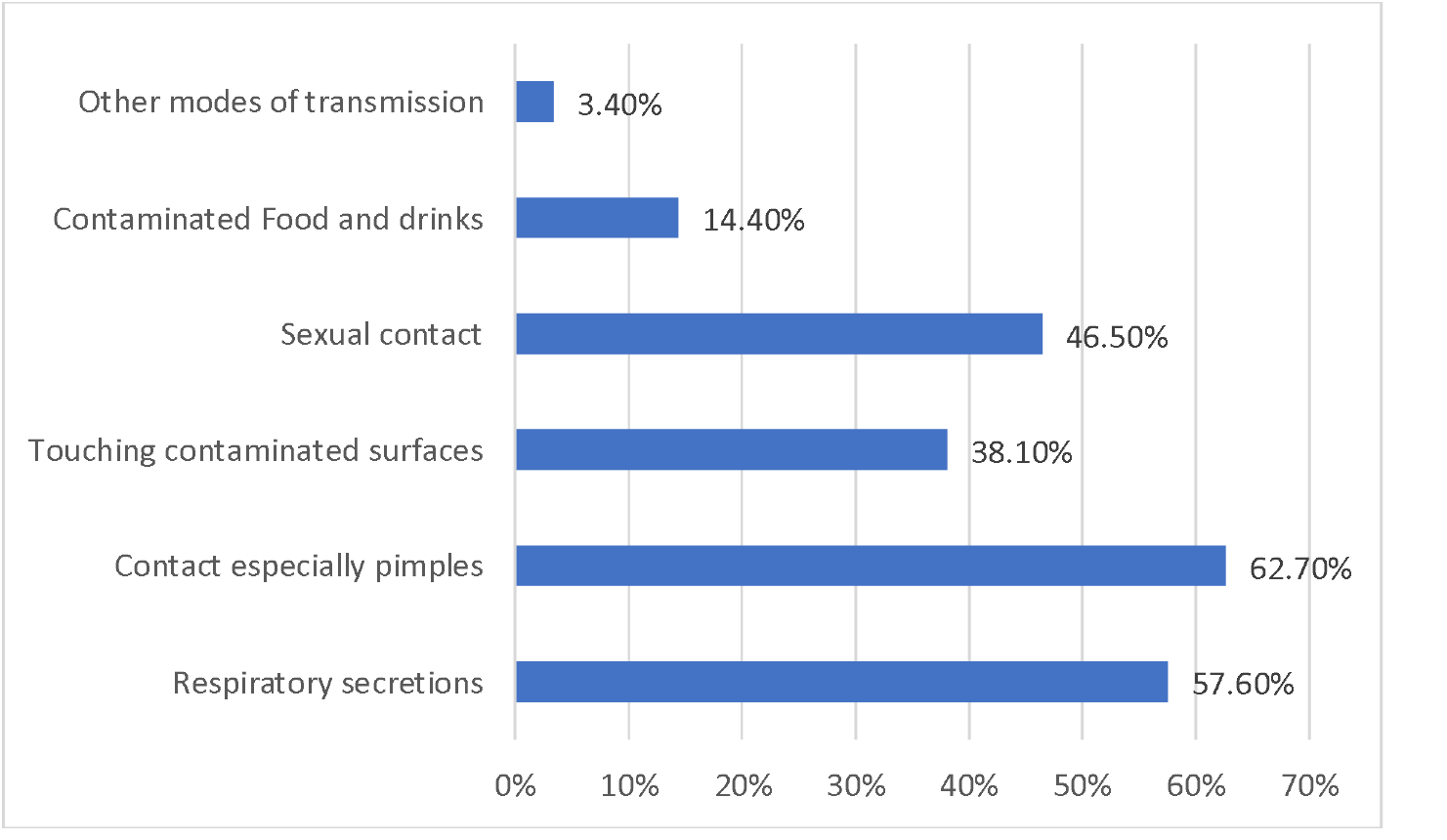
Public perception of Monkeypox disease transmission modes

Public participants’ overall mean Knowledge score of MPD was 4.88/9 points, SD= 1.50 points, highlighting a moderate level of knowledge. Knowledge score was dichotomized into low versus high based on the average score; the results showed that 56% measured relatively high (above average) MPD knowledge.

When considering the public participants’ sources of information about MPD, 45.6% relied on local official sources, including the MOH, 28% used International official health websites like Centers for Disease Control and Prevention (CDC) and WHO websites, but 66.1% relied on social networking and other internet-based sources. 61.3% of the participants perceived the need to seek more information about MPD.

To assess the variables associated with public participants’ high MPD knowledge scores, we ran a Multivariate Logistic Binary Regression Analysis. As shown in Table 1, age, employment status, and monthly household income did not correlate significantly with the odds of high knowledge score. However, respondents holding a university degree or other high educational levels were found to have significantly high scores (OR 1.35 p-value= 0.002). Respondents who previously developed COVID-19 disease had slightly but not significantly lower odds of a high score (OR .816 p-value=0.060). On the other hand, the respondents who had self and family worries about contracting MPD had significantly high knowledge scores (OR 1.843 p-value <0.001). All respondents’ sources of information correlated significantly with odds of high knowledge scores but with variable significance: local official sources, as MOH (OR 2.274, p-value <0.001), social media channels (OR 1.735, p-value <0.001), while official international sources, as the CDC and WHO (OR 1.583, p-value 0.001).

We assessed the public participants’ perceived need to seek more information about MPD after completing the current survey; 61.3% did show interest in seeking more information. Table 2 highlights the public’s characteristics associated with the perceived need to seek more information about MPD. Males were significantly less predicted to seek more information (42.2% times less, p=0.003). Age and educational level did not correlate significantly with their interest to seek more information, while the household’s income (HHI) >=15000 SR per month was associated significantly with less interest (9.6% times less, p-value=0.010). Participants who developed COVID-19 previously were found to be significantly less inclined to seek more information about MPD (39.8% times less, p-value=0.002).

**Table 2.** Public’s analysis of variables associated with high odds of seeking more information about Monkeypox.

|  | Multivariate<br>adjusted Odds<br>Ratio (OR) | 95% CI |  | p-value |
| --- | --- | --- | --- | --- |
|  |  | Lower | Upper |  |
| **Sex** |  |  |  |  |
| Male | 0.578 | 0.402 | 0.832 | .003 |
| **Age group** | 0.965 | .855 | 1.088 | .557 |
| **Educational Level** | 0.933 | .756 | 1.151 | .518 |
| **Households' Monthly income**<br>**15000 SR / Month** | 0.904 | 0.838 | 0.976 | .010 |
| **Previously developed COVID-19** | 0.602 | 0.435 | 0.832 | .002 |
| **Perceives Monkeypox as a**<br>**dangerous and virulent disease** | 1.559 | 1.243 | 1.954 | <0.001 |
| **Family compliance with universal**<br>**precautionary measures** | 1.093 | 1.002 | 1.192 | .044 |
| **Moderate or higher compliance** |  |  |  |  |
| **Monkeypox knowledge score** | 1.029 | .954 | 1.110 | .463 |
| **GAD7 score** | 0.997 | .976 | 1.018 | .772 |
| **Supports vaccination against MPD** | 1.674 | 1.338 | 2.095 | <0.001 |
| **Reliance on social networks and**<br>**nonofficial sources of information** | 0.418 | 0.247 | 0.709 | .001 |
| **Worried about a possible National**<br>**Lockdown due to MDP** | 1.438 | 1.099 | 1.881 | .008 |
| **Constant** | 1.736 |  |  | .138 |

DV= perception of the need to seek more information about MPD

However, those who perceived MPD as virulent and dangerous were significantly more inclined to seek more information (55.9% times more, p-value<0.001), in addition to those who were worried about possible national lockdown related to MPD (43.8% times more, p-value=0.008), echoing those who had moderate to high compliance with universal precautionary measures (9.3% times more, p-value=0.044). Participants’ MPD average knowledge score and GAD7 scores did not correlate significantly with their perceived need to seek more information. While those who supported vaccination against the disease were significantly more inclined to seek more information (67.4% times more, p-value<0.001). Use of social networks and other nonofficial sources of information were significant predictors of less inclined to seek more information about MPD (58.2% times less, p-value=0.001).

### HCWs’ MPD knowledge assessment

1130 HCWs participated in the survey. We assessed their MPD knowledge using four domains: vaccination, transmission modes, clinical presentation, and isolation precautions. Table 2 (Appendix) details the descriptive analysis of their responses. 61.9% of the participants correctly answered that it is caused by a Pox family virus. Regarding vaccination, only 28.3% correctly answered that (Jynneos) vaccine has dual activity against both smallpox and MPD. Interestingly, 79.7% incorrectly answered that (VARIVAX) vaccine, a chickenpox vaccine, is effective against MPD. While only 34.7% correctly answered that HCWs could benefit from post-exposure prophylaxis to MPD cases with the smallpox vaccine.

Regarding HWCs’ knowledge about MPD modes of transmission, nearly half correctly answered that animal-to-human transmission exists, most of them 64.8% had correctly agreed that human-human transmission via direct skin contact is a possible mode of transmission, 67.6% correctly answered that MPD is not an airborne transmissible disease. In comparison, only 53.7% correctly agreed it is transmitted via droplets. 94.4% correctly inferred that it is not a foodborne disease, and 96% believed falsely that other modes of transmission might indeed exist. Regarding the clinical presentation of MPD, only 57.5% correctly answered that it has similar symptoms to COVID-19 before the appearance of the rash. The majority correctly indicated that fever, rash, and headaches are indeed presenting symptoms of MPD. At the same time, less than half of them correctly inferred that myalgia and lymphadenopathy are MPD symptoms. But the majority > 90% of the participants had the misconception that respiratory distress, hemodynamic shock, seizures, and loss of smell sense, as well as acute kidney injury, are among the clinical manifestations of MPD. Regarding isolation precautions against MPD, 73.6% correctly inferred that contact precautions are needed, while 65.2% had incorrectly answered that airborne precautions might be needed in comparison to 53.8% who had correctly answered that droplet precautions are needed at a certain phase of the disease, while 96% incorrectly perceived that other isolation precautions are required to control the disease transmission.

### HCWs’ Monkeypox knowledge score

Table 3 displays the dissected HCWs’ knowledge score. The highest achieved score was of MPD precautionary modes score (mean score 2.9/4,72.5%, SD 0.8), followed by transmission modes (mean score was 6.2/9, 68.9%, SD 1.5). The overall HCWs’ MPD knowledge score was (mean 14.4/28, 51.6%, SD 3.8).

**Table 3:** HCWs’ knowledge and GAD7 scores.

|  | GAD7 |  |  |  |  |
| --- | --- | --- | --- | --- | --- |
|  | Mean score | Mean % score | SD | IQR | Maximum score |
| MPD transmission modes' knowledge score | 6.2 | 68.9 | 1.5 | 2.0 | 9 points |
| MPD vaccination knowledge score | 1.0 | 33.3 | 0.9 | 2.0 | 3 points |
| MPD clinical presentation knowledge score | 4.5 | 35.4 | 2.2 | 3.0 | 12 points |
| MPD precautionary methods knowledge score | 2.9 | 72.5 | 0.8 | 1.0 | 4 points |
| Overall MPD knowledge score | 14.4 | 51.6 | 3.8 | 5.0 | 28 points |

Public and HCWs’ MPD sources of information are displayed in figures 2 and 3. Most HCWs (74.2%) perceived they need to seek more information about MPD after receiving the survey. 57.6% used official local sources, 59.8% used official international health authorities’ sources, 51.1% used social media and internet-based networks, while 24.5% only relied on scientific Journals.

**Figure 2:**
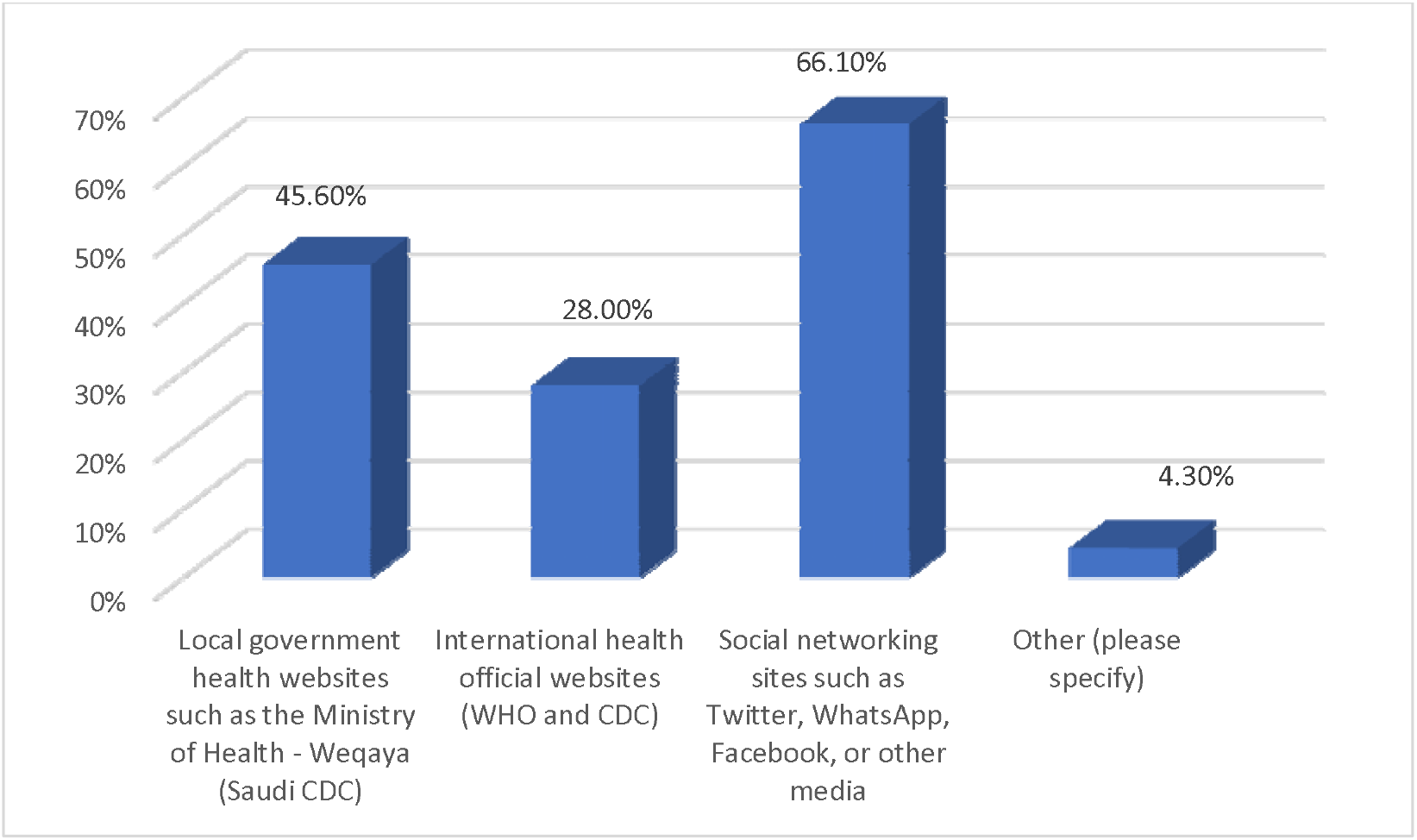
Public sources of information about Monkeypox disease

**Figure 3:**
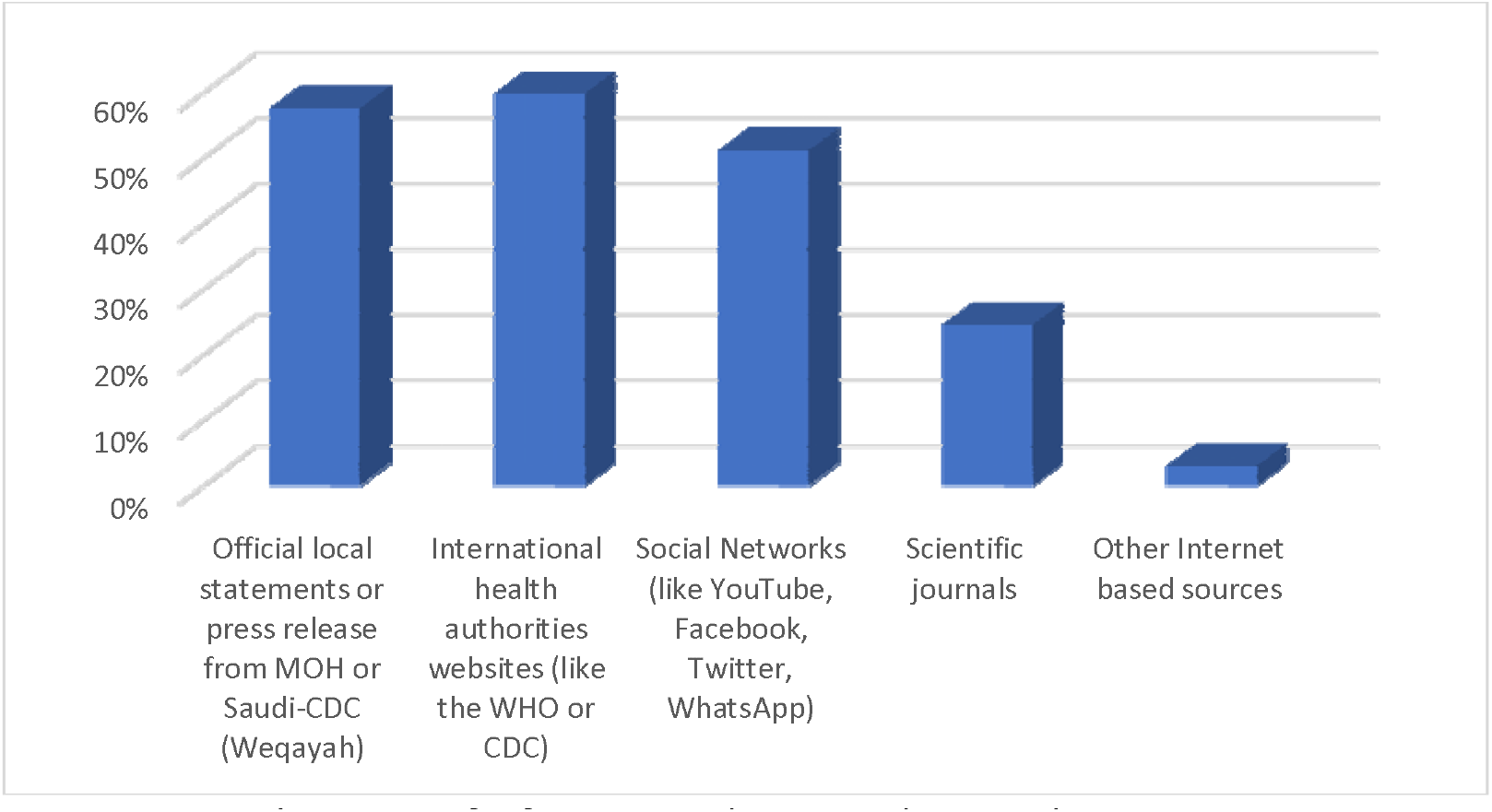
HCWs’ sources of information about Monkeypox disease

Table 4 shows variables associated with HCWs’ perceived need to seek more information about MPD after involvement in the current survey. HCWs’ sex, age, working hospital type, self-rated awareness level of MPD, participants’ GAD7, and overall MPD knowledge score did not correlate significantly with their odds to seek more information about the MPD.

**Table 4.**
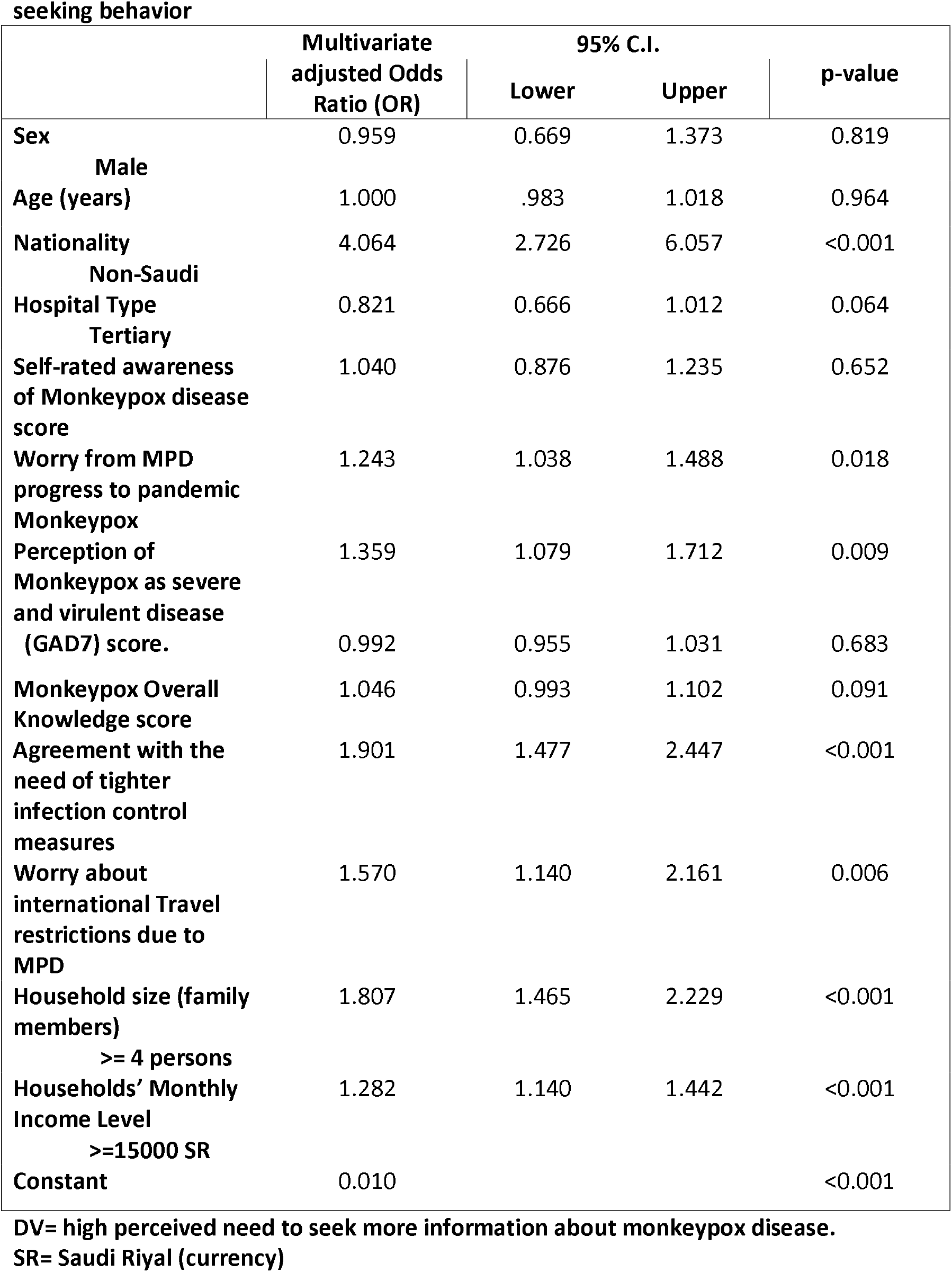
Multivariate Binary Logistic Regression analysis of the HCW’s MPD information-seeking behavior.

Expatriate HCWs had significant odds of seeking more information about the MPD (4.10 times more, p<0.001). HCWs’ worry about MPD progress to a pandemic was a significant positive predictor of their perceived need to read more about it (OR=1.243, p-value=0.018). Their worry about international travel restrictions due to the MPD was also a significant positive predictor (OR=1.57, p<0.001). Echoing that, their perception of MPD as a severe and virulent disease was another strong positive predictor (OR=1.359, p-value=0.009). The strongest predictor to seek more information about MPD was their perception of the need to apply tighter infection control prevention measures to control the progress of the disease (1.901 times more, p-value<0.001). HCWs with a household size of >=4 persons had a high perceived need to seek more information regarding MPD (OR=1.81, p<0.001), in addition to those who had monthly income >=15000 (OR=1.282, p<0.010).

To understand HCWs’ characteristics’ correlation with their assessed knowledge score, we ran a multivariate Linear Regression Analysis shown in Table 5. Male HCWs had significantly lower mean knowledge scores compared to females (β coefficient= −0.564, p-value=0.007), while the age did not correlate significantly with their knowledge score p=0.146. Physicians, compared to other HCWs, were found to be significantly more informed about the disease (β coefficient= 875 p<0.001). HCWs’ self-rated high awareness of MPD correlated positively and significantly with their knowledge score about it (β coefficient=0.741, p<0.001). Use of any source of information about MPD correlated significantly with higher knowledge score compared to using no source at all (β coefficient= 7.323, p<0.001). HCWs’ household monthly income correlated positively and significantly with their MPD knowledge score (β coefficient=0.133, p=0.045). Interestingly the HCWs’ perception of the need to seek more information about MPD did not correlate significantly with their knowledge score (p-value=0.098). Considering the participants’ working institution type, those who work in secondary and tertiary centres had significantly higher mean MPD knowledge scores (β coefficient= 0.700, 0.918, p-value 0.021, <0.001, respectively). While participants’ working in the operating rooms, outpatient department and general hospital wards had significantly lower mean MPD knowledge score compared to those working in other units (β coefficient= −1.135, −.696, −.564, p-values 0.047, 0.002, 0.016, respectively).

**Table 5.** Multivariate Linear Regression analysis of the HCW’s variable associated with their mean overall MPD Knowledge score.

|  | Unstandardized<br>Beta Coefficients | 95.0% CI for Beta<br>coefficient |  | p-value |
| --- | --- | --- | --- | --- |
|  |  | Lower<br>Bound | Upper<br>Bound |  |
| **(Constant)** | 4.716 | 3.471 | 5.961 | <0.001 |
| **Sex** |  |  |  |  |
| **Male** | -0.564 | -0.976 | -0.153 | 0.007 |
| **Age (years)** | 0.015 | -0.005 | 0.034 | 0.146 |
| **Clinical Role** |  |  |  |  |
| **Physicians** | 0.875 | 0.428 | 1.321 | <0.001 |
| **Self-rated awareness of**<br>**Monkeypox disease** | 0.741 | 0.565 | 0.916 | <0.001 |
| **Use of any source of**<br>**information** | 7.323 | 6.468 | 8.177 | <0.001 |
| **Households Monthly**<br>**Income** | 0.133 | 0.003 | 0.263 | 0.045 |
| **Perception of the need to**<br>**seek information about**<br>**MPD** | 0.402 | -0.074 | 0.878 | 0.098 |
| **GAD7 score** | -0.043 | -0.083 | -0.003 | 0.035 |
| **Hospital type** |  |  |  |  |
| **Secondary** | 0.700 | 0.107 | 1.293 | 0.021 |
| **Hospital type** |  |  |  |  |
| **Tertiary** | 0.918 | 0.446 | 1.390 | <0.001 |
| **Working unit** |  |  |  |  |
| **Operating Room** | -1.135 | -2.257 | -0.013 | 0.047 |
| **Working Unit** |  |  |  |  |
| **Outpatient area** | -0.696 | -1.138 | -0.254 | 0.002 |
| **Working unit** |  |  |  |  |
| **General hospital wards** | -0.564 | -1.022 | -0.107 | 0.016 |

**DV= Monkeypox disease overall Knowledge score. Model R=0.589, Adj. R-squared=0.34**

Table 6 shows the Spearman’s Rho test correlation between HCWs’ overall MPD knowledge score and their other cognitive MPD-related variables. The overall knowledge score correlated significantly and positively with their self-rated awareness of the MPD and their self-rated worry about MPD progress to a pandemic (rho=0.253, 0.260 p<0.010, p<0.050, respectively). On the other side, the HCWs’ GAD7 score correlated weakly and negatively with their MPD knowledge score, but positively and significantly with their worry about MPD progressing to a pandemic and their perception of it as severe and virulent disease (rho=.279, .200, p<0.010) respectively. The HCWs’ worry about MPD progress to a pandemic correlated significantly and positively with their perception of MPD as virulent and severe disease (rho=0.319, p<0.010).

**Table 6.**
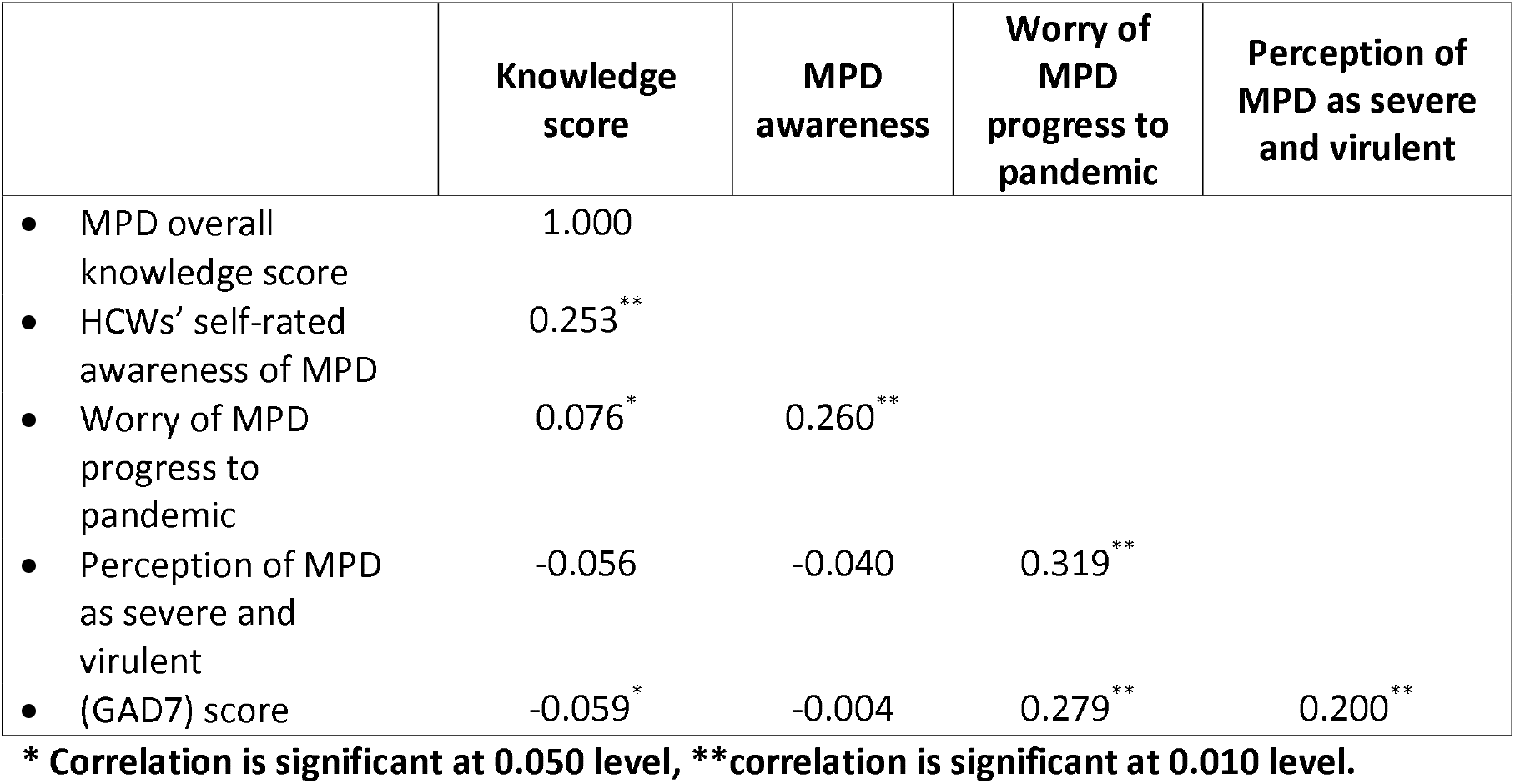
Bivariate Spearman’s (Rho) test correlations between HCWs’ MPD knowledge, awareness, perception of severity and worry about its progress to pandemic.

|  | Knowledge score | MPD awareness | Worry of MPD progress to pandemic | Perception of MPD as severe and virulent |
| --- | --- | --- | --- | --- |
| • MPD overall knowledge score | 1.000 |  |  |  |
| • HCWs' self-rated awareness of MPD | 0.253** |  |  |  |
| • Worry of MPD progress to pandemic | 0.076* | 0.260** |  |  |
| • Perception of MPD as severe and virulent | -0.056 | -0.040 | 0.319** |  |
| • (GAD7) score | -0.059* | -0.004 | 0.279** | 0.200** |

\* Correlation is significant at 0.050 level, \*\*correlation is significant at 0.010 level.

## Discussion

Knowledge dictates attitudes and practices, but compliance with knowledge is needed to assure achieving intended goals, in our case, dealing with infectious disease and preventive measures[21]. The literature on MPD still has gaps that remain critical to understanding and better informing healthcare stakeholders [22]. In this research, we aimed to assess the public and HCWs’ MPD knowledge, which is a newly re-emerging infectious disease that alerted humanity with multiple recent outbreaks just after its recovery from the harsh COVID-19 pandemic, while MPD has been considered from history or at least a rare disease.

Sources of knowledge affect its accountability, trust ability, and, therefore, its application. Knowledge of emerging infectious diseases is crucial in healthcare, especially in our case of MPD. While MPD used to be a zoonotic disease with limited transmissions, the sudden global rise of several human-human transmissions warrants careful attention. Despite detecting only three positive cases in the Saudi community, all coming from Europe[11], our sampled population from the HCWs and the public showed a moderate level of awareness about the disease even before these MPD were reported locally.

We found that both public and HCWs relied on local official health sources for about 45-57%. While HCWs used international official health sources in about 60% in comparison to the public who used it in 28%, which is explained by the HCWs’ educational and qualification basis, who also used scientific journals in 24.5% of the cases as a source of information. When considering social networks and other internet-based sources, the public used them more widely than HCWs (66% compared to 51%), which also stems from HCWs’ critical appraisal methodology of acquiring knowledge that deviates them from using sources with low accountability. A previous study on MPD in Africa reported that Media campaigns to raise awareness did not demonstrate effectiveness in changing public health behavior toward the disease[23]. Interestingly in our study, both groups had high rates of perceived need to seek more information about MPD, 61% for the public and 74% for HCWs, which indicates their high alert and concerns about this remerging disease.

We found major gaps in the HCWs’ knowledge about MPD, especially regarding vaccination, as the majority did not realize that (Jynneos) vaccine has dual activity against smallpox and (MPV). The vast majority falsely thought that (VARIVAX) vaccine, which is effective against chickenpox, is also effective against (MPV)[24–26].

Physicians and HCWs working in secondary and tertiary centres had odds of high MPD knowledge scores. Understandably, variations in knowledge would occur among HCWs; those working in secondary or tertiary centres potentially might have the richer knowledge base; this might be explained by their clinical exposure in their centres in addition to their higher expertise and qualification. While those working in the operating rooms, outpatient department and general hospital wards had significantly lower MPD knowledge scores. In a previous study in 2020, general practitioners who worked in community health centres had higher knowledge than those who worked in private clinics[24]. Another study showed that among HCWs in Italy, there was substantial knowledge gaps in relation to MPD[25]. Female HCWs and those who rated themselves with high self-awareness about the MPD had odds of high knowledge score, which translates to their female personality having augmented anxiety from illnesses in general compared to males and therefore tend to enrich their medical knowledge, especially that of preventive nature.

Among the general population, age, employment status, and monthly household income did not correlate with MPD knowledge scores. In contrast to this finding, a study from Saudi Arabia showed that these factors correlated positively with MPD level of knowledge in addition to participants’ marital status[26].

In relation to the public responses, variables associated with odds of high knowledge score partly echoed the HCWs’ variables, as the use of all different sources of information by the public and HCWs correlated significantly with the odds of high knowledge scores. Interestingly, those who perceived the need to seek more information about MPD did not have significant odds for either low or high knowledge scores, which reflects their perceived low knowledge base and need to enrich it. High public’ household monthly income was a predictor of a low public knowledge score, while it was a positive predictor for HCWs’ high knowledge score. As expected, the public holding university degree and higher educational levels scored high knowledge scores, which echoes our colleagues’ results[26]. Additionally, public participants’ who had self and family worry about contracting the disease had significantly high knowledge scores, which is again an expected intuitive healthy behaviour as worry dictates the perception of risk, which translates into preventive practice stemming from reading and acquiring targeted knowledge[27].

In addition, variables associated with odds of high knowledge score partly echoed the HCWs’ variables, as the different sources of information used by the public and HCWs correlated significantly with the odds of high knowledge scores. Interestingly, the tendency to seek more information about MPD did not correlate by any means in both groups with high knowledge scores. Public’ household monthly income surprisingly was not a correlating factor for public knowledge, while it was a positive predictor for HCWs’ high knowledge score. As expected, the public holding university degree and higher educational levels scored high knowledge scores, which echoes our colleagues’ results[26]. Additionally, public participants’ who had self and family worry about contracting the disease had significantly high knowledge scores, which is again an expected intuitive healthy behaviour as worry dictates the perception of risk, which translates into preventive practice stemming from reading and acquiring targeted knowledge[27].

Both Public and HCWs who showed interest to read more about the MPD overlapped by either having worry about the MPD progressing into a pandemic or national lockdown, or to cause international travelling restrictions. HCWs’ who perceived MPD as potentially severe and virulent and those who perceived the need to apply tighter infection control prevention measures to control the progress of the disease, perceived the demand to seek more information about MPD. Public almost mirrored HCWs’, as public participants who perceived MPD as virulent and dangerous, and those who had moderate to high compliance with universal precautionary measures also showed high interest to read more about MPD. Male public participants were less inclined to seek more information about MPD. Interestingly, those who already developed COVID-19 also showed less interest to seek more information, which can be explained by their feeling of passing through a universal pandemic safely, which can give reassuring credit through the current alert, as they probably have more optimism for the future[28].

On the other hand, this was not the case for public participants who supported vaccination against MPD, who showed significantly high interest to read more about the disease, reflecting their high alert level. Additionally, the public’s use of non-official sources of information about MPD was significantly associated with less incline to increase the MPD knowledge base; this might be explained by the impressive widespread application and usage of those sources as a source of information for patients and their family through support and e learning[29,30]

Our public participants’ correct answers regarding the modes of transmission of MPD ranged between 46-62%, even about 46% knew that sexual intimate relationships are a mode of transmission. Our results were largely consistent with that of another local questionnaire by Alshahrani et al., with 480 participants from the general population; most participants could correctly answer that it is transmitted through droplets (60%) and bodily fluids (53.8%) [26].

The knowledge gap and the incorrect answers could be attributed to the scarcity of cases within the community, and the lack of interest in pursuing more information regarding the MPD for some public. Another possible factor could be some misleading and untrustworthy social media reports and other internet resources, as most of our participants (66.1 %) considered them the primary reference. Although COVD-19 fatigue still overshadows international and national scenes, which could potentially make people less interested in acquiring information about the current MPD outbreak. Nonetheless, nearly two-thirds of public participants (61.3%) felt the need to seek more information about MPD.

Only 50% of HCWs correctly answered that animal-to-human transmission exists, with only 64.8% agreed that human-human transmission occurs. Interestingly, the HCW and the publics’ knowledge regarding the transmission mode was not vastly different. Consequently, most HCWs (73.6%) correctly concluded that standard contact precautions are needed, while 65.2% had incorrectly answered that airborne precautions might be needed, compared to 53.8% who had correctly agreed that droplet precautions might be needed at a certain phase of the disease. Similarly, an Italian study addressing HCWs’ knowledge showed that most participants recognized the potential transmission of MPD through the respiratory system via respiratory droplets, direct contacts, and body fluids, and that standard preventive measures may be sufficient to avoid infection (74.8%)[25].

Although the HCWs are at high risk of contracting the disease from symptomatic undiagnosed patients, a couple of hurdles could be identified as causes for the struggle to gain sufficient information regarding MPD. An influential factor is the limited awareness campaigns organized by the local and international health authorities like WHO aiming to familiarise the HCWs with the remerging disease, though limited research showed that previous MPD campaigns were less effective in raising awareness[23]. Other factors could be insufficient access to free, trustworthy scientific resources and, finally, the lack of time. Additionally, with many healthcare authorities still employing strict isolation precautions and infection control measures because of the COVID-19 pandemic, many HCWs might feel that they are taking sufficient precautions against other infectious diseases.

## Strengths and limitations

Our study is the first to explore the Saudi public and HCWs’ MPD knowledge in the early WHO MPD alert and assess interactions among them. We identified areas of improvement in both groups’ awareness and knowledge base for that emerging alerting infectious disease. We assessed knowledge related to different aspects vital for self-protection and to build efficient preventive strategies. Limitations of our results might be related to inability to dissect both groups based on their travelling experience and social activities. Other limitations stem from basic observational studies’ limitations in relation to sampling technique and being a knowledge assessment study; the responses are liable for recall bias.

## Conclusion

We found that both public and HCWs have a moderate level of MPD knowledge. Both had about 50% usage of social networks as sources of information, while still use of official health websites usage was comparable but higher for HCWs. HCWs had major gaps in their knowledge related to clinical presentation and vaccination against MPD. Both perceived need to seek more information about MPD did not converge with knowledge scores. Higher level of qualification, high disease awareness, and high worry of contracting the disease, were all associated with higher knowledge scores in both groups. While both interacted in the perceived need to seek more information for those who perceived MPD severe and virulent disease and the need to apply tighter infection control measures to control the current emerging outbreaks.

## Data Availability

Data is available from the corresponding author upon reasonable request.

## Abbreviations

CDC: Centers for Disease Control and Prevention
COVID-19: Coronavirus disease 2019
GAD7: Generalized Anxiety Disorder 7-item
HCW: Healthcare workers
MOH: Ministry of Health
MOXV: Monkeypox virus
MPD: Monkeypox disease
PHEIC: Public Health Emergency of International Concern
SARS-CoV-2: Severe acute respiratory syndrome coronavirus 2
WHO: World Health Organization

## Acknowledgments

The authors would like to thank Researchers Supporting Project number (RSP2022R507), King Saud University, Riyadh, Saudi Arabia for the financial support needed to conduct the study. The research team is thankful for the statistical data analysis consultation offered by www.hodhodata.com.

## Author contributions

MHT, FAL, SA, KA, and JA conceptualized the study, analyzed the data, and wrote the manuscript.

NA, SD, MH, RH, AA, AAH, SAS, FA, FAZ, ZM, and MB contributed to the study design; collected, analyzed, interpreted data; and edited the manuscript.

All authors reviewed and approved the final version of the manuscript.

## Appendix

**Appendix Table 1:** Publics’ Monkeypox knowledge assessment questions.

|  |  |  |
| --- | --- | --- |
| **Can Monkeypox be transmitted before symptoms appear, especially skin blisters?** | **No.** | **Frequency** |
| No | 793 | 51.3 |
| Yes | 753 | 48.7 |
| **Do you know if there is a cure for Monkeypox** |  |  |
| Unsure | 72 | 4.7 |
| There is no effective treatment, treatment is | 742 | 48 |
| There are antivirals to treat this disease | 732 | 47.3 |
| **Do you think that the Smallpox vaccine used before 1980 is effective against Monkeypox** |  |  |
| No | 108 | 7 |
| Unsure | 875 | 56.6 |
| Yes | 563 | 36.4 |
| **Do you think that the chickenpox vaccine is effective against Monkeypox** |  |  |
| No | 227 | 14.7 |
| Unsure | 1000 | 64.7 |
| Yes | 319 | 20.6 |
| **Overall Monkeypox Disease Knowledge score, mean (SD).**<br>**maximum possible score= 9** |  | 4.88 (1.49) |
| Low | 681 | 44 |
| High | 865 | 56 |
| **Perceived need to seek more information about Monkeypox disease** | 948 | 61.3 |

**Appendix Table 2:** HCWs’ MPD knowledge assessment questions analysis.

|  | Best correct answer | Incorrectly answered No. Frequency | Correctly answered No. Frequency |
| --- | --- | --- | --- |
| **Jynneos is an FDA-approved vaccine for the prevention of Monkeypox disease with dual activity against Smallpox and Monkeypox infections.** | True. | 808 (71.7) | 319 (28.3) |
| **Chickenpox vaccine, like Varivax, has dual activity against Chickenpox and Monkeypox infections.** | False. | 898 (79.7) | 229 (20.3) |
| **HCW exposed to Monkeypox case benefit from receiving post-exposure prophylaxis (PEP) with the smallpox vaccine** | True. | 736 (65.3) | 391 (34.7) |
| **Monkeypox is caused by the Pox family virus** | True | 429 (38.1) | 698 (61.9) |
| **Monkeypox disease transmission** |  |  |  |
| Animal to human | True. | 565 (50.1) | 562 (49.9) |
| Human to human via direct skin contact | True. | 397 (35.2) | 730 (64.8) |
| Human to human via sexual route | True. | 668 (59.3) | 459 (40.7) |
| Airborne | False. | 366 (32.5) | 761 (67.6) |
| Droplet | True. | 522 (46.2) | 605 (53.7) |
| Food-Borne | False. | 63 (5.6) | 1064 (94.4) |
| Contaminated water | False. | 98 (8.7) | 1029 (91.3) |
| Other modes | False. | 45 (4) | 1082 (96) |
| **Monkeypox disease presenting signs and symptoms** |  |  |  |
| Before the rash appears, the symptoms of COVID-19 and Monkeypox are very similar. | True. | 479 (42.5) | 648 (57.5) |
| Fever | True. | 186 (16.5) | 941 (83.5) |
| Rash | True. | 173 (15.4) | 954 (84.6) |
| Headaches | True. | 424 (37.6) | 703 (62.4) |
| Lymphadenopathy | True. | 599 (53.1) | 528 (46.9) |
| Myalgias | True. | 612 (54.3) | 515 (45.7) |
| Exhaustion-Physical | True. | 804 (71.3) | 323 (28.7) |
| Respiratory distress | False. | 265 (23.5) | 862 (76.5) |
| Shock-Hemodynamic instability | False. | 57 (5.1) | 1070 (94.9) |
| Seizures | False. | 57 (5.1) | 1070 (94.9) |
| Loss of the sense of smell | False. | 61 (5.4) | 1066 (94.6) |
| Acute kidney injury | False. | 47 (4.2) | 1080 (95.8) |
| **Monkeypox required isolation methods** |  |  |  |
| Contact precautions | True. | 298 (26.4) | 829 (73.6) |
| Airborne precautions | False. | 392 (34.8) | 735 (65.2) |
| Droplet precautions | True. | 521 (46.2) | 606 (53.8) |
| Other isolation techniques | False. | 24 (2.1) | 1103 (97.9) |

## References

1. CDC. 2022 Monkeypox Outbreak Global Map. Available online: https://www.cdc.gov/poxvirus/monkeypox/response/2022/world-map.html (accessed on 13 Jul 2022).

2. Strassburg, M.A. The global eradication of smallpox. Am J Infect Control 1982, 10, 53–59, doi:10.1016/0196-6553(82)90003-7.

3. Simpson, K.; Heymann, D.; Brown, C.S.; Edmunds, W.J.; Elsgaard, J.; Fine, P.; Hochrein, H.; Hoff, N.A.; Green, A.; Ihekweazu, C.; et al. Human monkeypox - After 40 years, an unintended consequence of smallpox eradication. Vaccine 2020, 38, 5077–5081, doi:10.1016/j.vaccine.2020.04.062.

4. Thornhill, J.P.; Barkati, S.; Walmsley, S.; Rockstroh, J.; Antinori, A.; Harrison, L.B.; Palich, R.; Nori, A.; Reeves, I.; Habibi, M.S.; et al. Monkeypox Virus Infection in Humans across 16 Countries - April-June 2022. N Engl J Med 2022, doi:10.1056/NEJMoa2207323.

5. WHO. Monkeypox Questions and Answers. Available online: https://www.who.int/news-room/questions-and-answers/item/monkeypox?%20gclid=EAIaIQobChMI7umqiYi8-AIVE-3tCh0E2AAgEAMYASAAEgLzJPD_BwE (accessed on 16 Oct 2022).

6. Antinori, A.; Mazzotta, V.; Vita, S.; Carletti, F.; Tacconi, D.; Lapini, L.E.; D’Abramo, A.; Cicalini, S.; Lapa, D.; Pittalis, S.; et al. Epidemiological, clinical and virological characteristics of four cases of monkeypox support transmission through sexual contact, Italy, May 2022. Euro Surveill 2022, 27, doi:10.2807/1560-7917.es.2022.27.22.2200421.

7. Peiró-Mestres, A.; Fuertes, I.; Camprubí-Ferrer, D.; Marcos, M.; Vilella, A.; Navarro, M.; Rodriguez-Elena, L.; Riera, J.; Català, A.; Martínez, M.J.; et al. Frequent detection of monkeypox virus DNA in saliva, semen, and other clinical samples from 12 patients, Barcelona, Spain, May to June 2022. Euro Surveill 2022, 27, doi:10.2807/1560-7917.es.2022.27.28.2200503.

8. WHO. Multi-country monkeypox outbreak: situation update. Available online: https://www.who.int/emergencies/disease-outbreak-news/item/2022-DON396 (accessed on 13 Jul 2022).

9. WHO. WHO Director-General declares the ongoing monkeypox outbreak a Public Health Emergency of International Concern. Available online: https://www.who.int/europe/news/item/23-07-2022-who-director-general-declares-the-ongoing-monkeypox-outbreak-a-public-health-event-of-international-concern (accessed on 10 Oct 2022).

10. Al-Tawfiq, J.A.; Kattan, R.F.; Memish, Z.A. Mass Gatherings and Emerging Infectious Diseases: Monkeypox is the Newest Challenge. In J Epidemiol Glob Health; 2022; Volume 12, pp. 215–218.

11. Agency, S.P. Health Ministry: First Monkeypox Case Reported in Saudi Arabia Available online: www.spa.gov.sa/2370035 (accessed on 10 Oct 2022).

12. Temsah, M.H.; Alhuzaimi, A.N.; Alamro, N.; Alrabiaah, A.; Al-Sohime, F.; Alhasan, K.; Kari, J.A.; Almaghlouth, I.; Aljamaan, F.; Al Amri, M.; et al. Knowledge, Attitudes, and Practices of Healthcare Workers During the Early COVID-19 Pandemic in a Main, Academic Tertiary Care Centre in Saudi Arabia. Epidemiol Infect 2020, 1-29, doi:10.1017/S0950268820001958.

13. Temsah, M.H.; Al-Sohime, F.; Alamro, N.; Al-Eyadhy, A.; Al-Hasan, K.; Jamal, A.; Al-Maglouth, I.; Aljamaan, F.; Al Amri, M.; Barry, M.; et al. The psychological impact of COVID-19 pandemic on health care workers in a MERS-CoV endemic country. J Infect Public Health 2020, 13, 877–882, doi:10.1016/j.jiph.2020.05.021.

14. Temsah, M.H.; Barry, M.; Aljamaan, F.; Alhuzaimi, A.N.; Al-Eyadhy, A.; Saddik, B.; Alsohime, F.; Alhaboob, A.; Alhasan, K.; Alaraj, A.; et al. SARS-CoV-2 B.1.1.7 UK Variant of Concern Lineage-Related Perceptions, COVID-19 Vaccine Acceptance and Travel Worry Among Healthcare Workers. Front Public Health 2021, 9, 686958, doi:10.3389/fpubh.2021.686958.

15. Barry, M.; Temsah, M.-H.; Aljamaan, F.; Saddik, B.; Al-Eyadhy, A.; Alanazi, S.; Alamro, N.; Alhuzaimi, A.; Alhaboob, A.; Alsohime, F. COVID-19 vaccine uptake among healthcare workers in the fourth country to authorize BNT162b2 during the first month of rollout. medRxiv 2021.

16. Alhasan, K.; Aljamaan, F.; Temsah, M.H.; Alshahrani, F.; Bassrawi, R.; Alhaboob, A.; Assiri, R.; Alenezi, S.; Alaraj, A.; Alhomoudi, R.I.; et al. COVID-19 Delta Variant: Perceptions, Worries, and Vaccine-Booster Acceptability among Healthcare Workers. Healthcare (Basel) 2021, 9, doi:10.3390/healthcare9111566.

17. Ajman, F.; Alenezi, S.; Alhasan, K.; Saddik, B.; Alhaboob, A.; Altawil, E.S.; Alshahrani, F.; Alrabiaah, A.; Alaraj, A.; Alkriadees, K.; et al. Healthcare Workers’ Worries and Monkeypox Vaccine Advocacy during the First Month of the WHO Monkeypox Alert: Cross-Sectional Survey in Saudi Arabia. Vaccines (Basel) 2022, 10, doi:10.3390/vaccines10091408.

18. Temsah, M.H.; Aljamaan, F.; Alenezi, S.; Alhasan, K.; Saddik, B.; Al-Barag, A.; Alhaboob, A.; Bahabri, N.; Alshahrani, F.; Alrabiaah, A.; et al. Monkeypox caused less worry than COVID-19 among the general population during the first month of the WHO Monkeypox alert: Experience from Saudi Arabia. Travel Med Infect Dis 2022, 49, 102426, doi:10.1016/j.tmaid.2022.102426.

19. Spitzer, R.L.; Kroenke, K.; Williams, J.B.; Lowe, B. A brief measure for assessing generalized anxiety disorder: the GAD-7. Arch Intern Med 2006, 166, 1092–1097, doi:10.1001/archinte.166.10.1092.

20. AlHadi, A.N.; AlAteeq, D.A.; Al-Sharif, E.; Bawazeer, H.M.; Alanazi, H.; AlShomrani, A.T.; Shuqdar, R.M.; AlOwaybil, R. An arabic translation, reliability, and validation of Patient Health Questionnaire in a Saudi sample. Ann Gen Psychiatry 2017, 16, 32, doi:10.1186/s12991-017-0155-1.

21. Hammerschmidt, J.; Manser, T. Nurses’ knowledge, behaviour and compliance concerning hand hygiene in nursing homes: a cross-sectional mixed-methods study. BMC Health Serv Res 2019, 19, 547, doi:10.1186/s12913-019-4347-z.

22. Kipkorir, V.; Dhali, A.; Srichawla, B.; Kutikuppala, S.; Cox, M.; Ochieng, D.; Nyaanga, F.; Găman, M.A. The re-emerging monkeypox disease. Trop Med Int Health 2022, doi:10.1111/tmi.13821.

23. Wogu, J.O.; Chukwu, C.O.; Orekyeh, E.S.S.; Nwankiti, C.O.; Okoye-Ugwu, S. Assessment of media reportage of monkeypox in southern Nigeria. Medicine (Baltimore) 2020, 99, e17985, doi:10.1097/md.0000000000017985.

24. Harapan, H.; Setiawan, A.M.; Yufika, A.; Anwar, S.; Wahyuni, S.; Asrizal, F.W.; Sufri, M.R.; Putra, R.P.; Wijayanti, N.P.; Salwiyadi, S.; et al. Knowledge of human monkeypox viral infection among general practitioners: a cross-sectional study in Indonesia. Pathog Glob Health 2020, 114, 68–75, doi:10.1080/20477724.2020.1743037.

25. Riccò, M.; Ferraro, P.; Camisa, V.; Satta, E.; Zaniboni, A.; Ranzieri, S.; Baldassarre, A.; Zaffina, S.; Marchesi, F. When a Neglected Tropical Disease Goes Global: Knowledge, Attitudes and Practices of Italian Physicians towards Monkeypox, Preliminary Results. Trop Med Infect Dis 2022, 7, doi:10.3390/tropicalmed7070135.

26. Alshahrani, N.Z.; Alzahrani, F.; Alarifi, A.M.; Algethami, M.R.; Alhumam, M.N.; Ayied, H.A.M.; Awan, A.Z.; Almutairi, A.F.; Bamakhrama, S.A.; Almushari, B.S.; et al. Assessment of Knowledge of Monkeypox Viral Infection among the General Population in Saudi Arabia. Pathogens 2022, 11, doi:10.3390/pathogens11080904.

27. Prati, G.; Pietrantoni, L. Knowledge, Risk Perceptions, and Xenophobic Attitudes: Evidence from Italy During the Ebola Outbreak. Risk Anal 2016, 36, 2000–2010, doi:10.1111/risa.12537.

28. Hodkinson, B.; Gina, P.; Schneider, M. New life after near death: Surviving critical COVID-19 infection. Afr J Thorac Crit Care Med 2021, 27, doi:10.7196/AJTCCM.2021.v27i4.184.

29. Langford, A.T.; Orellana, K.T.; Buderer, N. Use of YouTube to watch health-related videos and participation in online support groups among US adults with heart disease, diabetes, and hypertension. Digit Health 2022, 8, 20552076221118822, doi:10.1177/20552076221118822.

30. Klein, A.Z.; Magge, A.; O’Connor, K.; Gonzalez-Hernandez, G. Automatically Identifying Twitter Users for Interventions to Support Dementia Family Caregivers: Annotated Data Set and Benchmark Classification Models. JMIR Aging 2022, 5, e39547, doi:10.2196/39547.

